# Rolling with the F-words Life Wheel: reflexive thematic analysis of a coaching-based, holistic approach to pediatric occupational therapy

**DOI:** 10.1101/2023.03.04.23286803

**Authors:** Khushi Sehajpal, Claire McCrostie, Lucy Charles, Arul Hamill, Pio Terei, James Hamill

## Abstract

**Background and purpose:** The F-words Life Wheel approaches child development by hybridizing a holistic model in the F-words for Child Development, and a coaching model in Occupational Performance Coaching, along with a life-flow approach in the Kawa model. The effect of the F-words Life Wheel has not been previously studied. The purpose of this paper is to report parents’ experiences with the F-words Life Wheel.

**Methods:** This was a qualitative study based on interviews with parents of children with developmental needs and experts in child development. Interviews were conducted in person and transcribed verbatim. The researchers used reflexive thematic analysis within a critical realist paradigm.

**Results:** A total of 13 interviews were conducted, 11 with parents of children with developmental needs and two with child development experts. Interview transcripts totaled 42,763 words from which we developed 45 codes and three themes. The themes were 1) overwhelming, 2) power rebalance, and 3) connectedness. The overwhelming theme addresses how life with developmental needs is challenging, engaging with the health and disability system is difficult, and the focus on deficits can lead to a sense of being overwhelmed. The power rebalance theme addresses the transition from professionals calling the shots to giving agency to the child and family. Holistic goal setting empowers parents and children to direct and prioritize therapy, and helps shift from a deficit-focused to a “can-do” attitude. The connectedness theme addresses the linkages between psychological health, physical health, the extended family, and the planet as a whole.

**Conclusions:** The F-words Life Wheel approach appears to be empowering and motivating for children and families. Further research is needed to explore how holistic models of therapy such as the F-words Life Wheel can promote family-centered care and connectedness on a wider scale.

## Introduction

Childhood disability refers to any health-related condition that currently or in the future may limit a child’s capacity to adequately develop and participate in society (Halfon et al., 2012). Disability encompasses physical impairments as well as emotional, neurological, and behavioral problems. The biomedical model of medicine, which focuses on symptom profiles and measurable biological variables, is not always the best approach to child disability (Farre & Rapley, 2017). Recognizing the limitations of the biomedical model, Engel (1977) described the biopsychosocial model that encompasses psychological, social, environmental, and any societal complementary systems in place for individuals in the broader, systems approach to disease and healthcare (Engel, 1977, 1981). Building on the biopsychosocial model, the World Health Organization developed the International Classification of Functioning, Disability and Health (ICF) (WHO, 2002). The ICF framework views the health condition, body structure and function, activity limitations, participation restrictions, personal factors, and the environment in a dynamic interaction.

To facilitate the use of the ICF framework for children, Rosenbaum and Gorter developed the “F-words for Child Development” (Rosenbaum, 2022; Rosenbaum & Gorter, 2012). The six “F-words” − function, family, fitness, fun, friends, and future − targeted key areas of a child’s development. The catchy name acted as an aide mémoire, helped eliminate jargon, and made the ICF more accessible to families by contextualizing it in a way that was familiar and understandable. Developed in partnership with CanChild in Canada, the F-words were general enough to be applied globally (Soper et al., 2019) and have now been translated into over 30 languages (Cross et al., 2022).

In Aotearoa New Zealand, Charles et al. built on the F-words for Child Disability with the F-words Life Wheel (FWLW). In using the FWLW, therapists assist children and parents to rate each F-words domain on a radar chart or wheel (a process that can be repeated at intervals), set goals using the Canadian Model of Occupational Performance and Engagement (E. A. Townsend & Polatajko, 2007), work towards goals using the Occupational Performance Coaching approach (Graham, 2020), and journey in life using professional coaching and using the metaphor of a river as described by the Kawa model (Teoh & Iwama, 2015).

The purpose of this study was to explore the experiences of families of children with disability, and to describe their experiences with the FWLW. Being situated in Aotearoa New Zealand, a specific purpose was to explore Māori health models in relation to the holistic approach to child development. Therefore, our research questions were: 1) what are the lived experiences of parents of children with developmental needs, 2) how had the FWLW affected their child’s therapy, and 3) how could other models of health and wellness inform therapy for children with developmental needs.

## Methodology

The qualitative methodology used for this research was reflexive thematic analysis (Braun & Clarke, 2006, 2019, 2021b), a well documented approach that suited our research team’s range of experience with qualitative research, and a flexible approach that allowed us to interpret participants’ dialogues as realities (experiential) (Braun & Clarke, 2021a) while also viewing data through the lens of pre-existing frameworks, in keeping with the aims of the study. Reflexive thematic analysis embraces the researcher’s subjectivity and reflections. The theoretical framework was one of critical realism (Bhaskar, 2013; Yucel, 2018) which, in the words of Pilgrim, “rebalances our focus back onto being rather than knowing, without losing sight of epistemological matters” (Pilgrim, 2014). Critical realism recognizes that research is based in theory, that “reality is deep”, and that “the enquirer is part of the object of their enquiry” (Pilgrim, 2014). This suited our study of a practical, hand-on approach to child therapy.

The two researchers who interviewed participants (KS and CM) were fresh to the field of child development and had no prior experience with the FWLW; KS was a biomedical science post-graduate and the CM was a medical student. Two researchers were experienced pediatric occupational therapists (LC and AH). They had developed the FWLW in collaboration with CanChild. Most of the participants in the study were their clients. They were involved in the concept and design of the study and in the formulation of implications but, because of their close relationship with participants and the instrument being discussed, did not perform interviews or undertake the thematic analysis. Another researcher had worked with LC and AH on exploring how the FWLW could be interpreted in the light of Māori culture (PT). This researcher had many years’ experience in youth advocacy and was an expert in Māoritanga (practices, beliefs and culture of the Māori people). The final researcher was a doctor in a children’s hospital (JH).

The setting of the study was based around a community pediatric therapy practice. The context was a group practice of occupational therapists and physiotherapists in Auckland, Aotearoa New Zealand.

A purposive sampling strategy was used. Two authors, AH and LC, identified potential participants from amongst families who had worked with the FWLW in order to gain insights into the experiences of families with developmental diversity. One author (JH) identified potential participants from amongst pediatric colleagues in order to gain perspectives from professionals who had experience in both child development and Māori culture.

Sample size was determined pragmatically by the number of participants available during the period of the summer studentship. Retrospective appraisal of the sample size was guided by the concepts espoused by Braun and Clarke for thematic analysis (Braun & Clarke, 2021b). The concept of “saturation” was not appropriate for our qualitative approach and theoretical paradigm. Instead we used the concept of “information power” (Malterud et al., 2016). Our considerations in determining the information power of our sample were: 1) the aim of this study was relatively narrow, that is to hear the experiences of families with developmentally challenged children specific to the FWLW, justifying a smaller sample; 2) the specificity of the sample was dense because we used purposive sampling to invite a set of participants with experiences specific to our aims, favoring a smaller sample; 3) several theoretical frameworks underlie the FWLW (for example, the ICF framework, Occupational Performance Coaching, the Kawa Model); however, the FWLW itself does not represent an established theory − this would favor a larger sample; 4) the quality of the dialogue in our interviews was strong, favoring a smaller sample; and 5) the analysis was case-based, favoring a smaller sample. Based on these five criteria, a sample of 13 participants seemed reasonable.

This study was approved by the Auckland Health Research Ethics Committee (AH23374). Formal written consent was obtained for each participant through an online survey after providing background about the study and a participant information sheet. Additional verbal consent was obtained prior to the beginning of each interview.

Data were collected from transcribed interviews. Two researchers, KS and CM, together interviewed all participants either face-to-face, on a videoconferencing platform, or over the phone. Participants informed interviewees prior to the interview of which they would prefer. Interviews were semistructured, allowing participants to tell their story but also guided as shown in Appendix 1. Interviews lasted 40 – 60 minutes. Interviews were performed between December 2021 and April 2022.

Interviews were recorded on the videoconferencing platform or using the recording function on a phone. Two researchers (KS and CM) transcribed all interviews except for two that were transcribed by a research assistant (MM − see acknowledgments). Transcripts were serially numbered and deidentified. We assigned a randomly generated (using random.org) two-letter code to each interviewee for the purpose of reporting quotations in this paper. Transcripts were emailed to participants for verification and correction. Transcripts were securely stored on a password protected cloud platform managed by the University of Auckland. Data coding was performed on Taguette (Rampin & Rampin, 2021).

We used the process of reflexive thematic analysis described by Braun and Clarke (Braun & Clarke, 2022). Coding and theme development was a recursive process involving immersion in data, review of relevant literature, and reflection. Two researchers coded the data separately and met regularly to discuss and mold themes. We coded mainly inductively because it allowed for identification of patterns from participants’ experiences with child disability. However, we recognize the influence of the ICF framework, hauora, and other models on our mind-frame as we coded. Our interest in these models as well as participants’ experiences meant that theme development was both semantic and latent. We aimed to describe and interpret the experience of families with disability and with the FWLW at the semantic level as well as look for latent meaning related to the holistic models.

Although we took a critical realist approach, the “critical” side was almost constructionist in our consideration of power dynamics in society in general and healthcare in particular, and in our consideration of connectedness in the context of traditional ecological knowledge.

Researchers read transcripts twice to become familiar with the data and with what was said. We isolated quotations from the data and organized them under code labels. Codes were analyzed for similarity and organized into initial themes. Initial themes were refined and reviewed to ensure relevance to the research question and shared meaning between participants. We drew mind maps to help visualize the diversity and connectedness of themes as they developed. Themes were given informative names to reflect their scope and focus (Braun & Clarke, 2021). It should be noted that this was a recursive process and new codes, code labels, and themes continued to be generated even at later stages of analysis.

Two researchers, KS and JH, independently coded data and developed themes. The two researchers conferred regularly during the process of developing and refining themes. Themes were then checked with a third interviewer (CM). Following this, the two occupational therapist researchers who were considered experts in the FWLW (LC and AH) looked over the analysis and met with the rest of the team to offer modifications and enhancements to theme development and understanding. Next, through discussion with the author with expertise in Māori culture (PT), the research team further refined themes. A researcher with training in psychology read and offered advice from a psychology background.

To enhance trustworthiness of this report we took a lead from Braun and Clarke’s guidance on quality and reporting (Braun & Clarke, 2021c) and from the guidelines for publication of qualitative research by Elliot *et al*.*(Elliott et al*., *1999)* First we applied Braun and Clarke’s 15-point checklist of criteria for good thematic analysis to our study (Braun & Clarke, 2006) as shown in Appendix 2. Next, we used Nelson’s conceptual depth scale to self evaluate robustness of our conceptual categories (Nelson, 2017) as shown in Appendix 3. Guided by Tracy’s eight criteria for excellent qualitative research (Tracy, 2010) we applied triangulation and crystallization through multiple layers of credibility checks. Members of the research team brought a range of backgrounds and perspectives to the analysis which contributed to “triangulation” (validating theme accuracy) as well as “crystallization” (deepening understanding and complexity).

## Results

We interviewed 13 participants in total. Eleven participants were parents of children with developmental needs and two were health professionals who had expertise in child and youth health and in Māori health models. Eleven were female and two male. Three identified as Māori and eight as European. Six were in the 35−44 year age bracket, two were younger than 35 years and three older than 44 years.

Transcripts of the interviews amounted to 42,763 words. From these, we coded 170 extracts into 45 codes. Through an iterative process of reviewing, merging, mapping, and renaming we developed three key themes as shown in Table 1. The three themes were: “overwhelming,” “power rebalance,” and “connectedness.”

**Table 1.** Themes developed from the interviews.

| Theme | Description |
| --- | --- |
| Overwhelming | For both parents and children, life with developmental challenges is really hard, draining, and can be overwhelming. Engaging with the health and disability system is difficult. Focus on deficits adds to the sense of overwhelming. |
| Power rebalance | Transition from professionals calling the shots to giving agency to the child and family. Holistic goal setting empowers parents and children to direct and prioritize therapy. Shift from deficit-focused to “can-do” attitude. |
| Connectedness | Psychological health and physical health are linked along with the extended family and the planet. Connectedness of mind and body. Connectedness of the individual, family, and humanity. |

### Theme 1: Overwhelming

Challenges in navigating life with disability was a prevalent theme though many of the interviews. Parents described their experiences as overwhelming, draining, never doing enough, reserves depleted, capacity limited, always the “what’s wrong”.

> So before we learnt about the F-words, we had − it was really hard. I’ve had all these therapists and teachers and people all with their own ideas − “oh we should do this, we should do that” − and so then I was always trying to accommodate their requirements and go to all these appointments and it was all just like completely overwhelming all the time. (EQ)
>
> You sort of get thrown into quite a medical wheel; and so it does feel a bit like that, like the hamster on the wheel thing − actually rolling around and being seen by so many different people and not really having control. (GT)

The above extracts from two participants show the depth of personal strain resulting from multiple healthcare providers imposing each of their professional “prescriptions” on families. The hamster on the wheel metaphor invokes not only running off to multiple appointments, but also gives a sense of being directionless. Accommodating the requirements of health professionals calls into question who therapy centers on. The “medical wheel” metaphor invokes rotation around the health professional.

> It was just overwhelming, like I thought I was always on catchup. I was always trying to do everything that came across our path to help her. We had lots of appointments and we couldn’t see all the little things that we were doing that were so beneficial, if that makes sense. (EQ)
>
> But I think that our capacity is generally just a lot more limited, because so much time and effort goes into the child or children. And then you’re − yeah − guess your reserves, your reserves are a bit depleted, and you don’t always think about it. (GT)

Here, parents expressed how busy and draining life was for them in the medical model. Exhaustion is exacerbated by a feeling of failure:

> … whereas before I never thought I was doing enough. And then constantly overwhelmed… I felt like I was failing at everything. Whereas now I feel like we’re doing so well. (EQ)

A major contribution to exhaustion came from the deficit-focused system.

> And then you’re like, oh, goodness, it’s like, yeah, this bit went wrong, and this bit’s wrong, and then this happened, and then this happens… and then these things that aren’t going right. (GT)
>
> Being a parent of a child with special needs and going through all the extra medical side that is required and all the additional checks and all the additional therapy, and then getting them into kindi [Kindergarten] and then transitioning them into mainstream school, and then along with that you’ve got government requirements and… requirements for funding etcetera. So it is very deficit focused. You sit there and it’s like, “what can he not do.” So what does that mean for support? (YT)

Service providers − therapists, doctors, early learning, school, funders − magnified the child’s deficits each from their own perspective. This changed with the FWLW.

> Well, I think the biggest difference is that it has changed the mindset because it used to always be about the deficits and what you couldn’t do. (AL)

### Theme 2: Rebalancing Power

The FWLW lent a positive, can-do approach to therapy. Goal-setting empowered families to set boundaries, priorities, and to feel more comfortable with saying “no” to certain clinical goals, thus increasing their sense of autonomy. Families were more motivated to achieve their own goals, in contrast to the benchmarks that children were expected to achieve at certain ages, benchmarks that often left them feeling disheartened. Instead, children were given the power to say “I can do it.”

> … and the F-words reframes it to all about what you can do and what you’d like to do and how you need to get there. So it’s a much more positive way of thinking, which is so much better for self-esteem and confidence and all that sort of thing. (AL)
>
> The F-words Life Wheel is going, “hey I can do it, it will take me longer and I’ll make a bit of a mess but I’m doing it.” (YT)
>
> And I was like, “yeah we could but it doesn’t align with her goals, it’s not really a problem, we can’t do everything at once, so no, you know, it’s a good idea but it’s just not a priority at the moment”… It’s a good way to weed things out and not trying to overdo it all the time. (EQ)
>
> Once you fill it [the F-words Life Wheel] out you can see where the gaps are, what direction to go in and what’s important to get more support in. It’s just about a balance, but in order to succeed it needs to have the child’s input because they need to have the execution to do the goals and the plan and be successful. (AL)
>
> The F-word wheel sort of demands that parents are included so you can’t do one without including the parent. And that’s when you [directed to their child] started being included more. (AL)

Empowerment was limited by providers continuing to work with a different mental model from the holistic, strengths-based approach of the FWLW. It was quite common for parents to be given contradictory advice by different providers. Centering control with the family was seen as one way to counter the disconnectedness of services. Another suggestion from some families was to try to make the use of the FWLW more widespread across providers.

> I find that the public OT [Occupational Therapist] will say something different to the private OT and the public physio will say something different to the private physio and there are heaps of stakeholders and they are all saying sort of different things. Sometimes they are on the same page and sometimes they are not. (AL)
>
> The improvement would be – there are two parts to it − it being used consistently in different areas, in health and education, in [my child’s] situations at least. I don’t know whether this is something that is adopted in those areas for children, or children with disabilities, but it would be good if we all had the same template to work from. In education, I don’t think they use F-words as a standard structure – they have got their own sort of education structure, but that would be an improvement. And that’s more about utilizing it consistently across the various agencies that [my child] deals with. (QC)
>
> Well, it would be kind of useful if it was probably more widespread. So you know if this was at least available as a resource to like the Ministry of Education when they are going through like the IEPs [individual educational plan]. (YT)
>
> And then I think also because it is like a mind shift for all the therapists and educators and you know the medical community as well. To like shift the focus so they are not just deficit looking, it’s like being able to look at the whole child and the whole life. (YT)
>
> It would be its use across the board. The more familiar it is in the world of disability, the more you would use it and the more valuable it would be. If it was a common language across all aspects of our care then that would be really valuable. (QA)

### Theme 3: Mental well-being and connectedness

Extending from the empowerment theme was the deeper sense of self, mental health, and connectedness that came though in many interviews. Connectedness went beyond the usual therapy domains of physical and intellectual capabilities, and was expressed in terms of spirituality, faith, and mana (authority, power, prestige (*Paekupu*, 2022)).

> For us, spirituality would be focused on the connection, I think. Connection with people and the environment. It would be for my child, fostering her self-confidence and her identity as someone having a disability. (EQ)
>
> … a recognition that well-being encapsulates all aspects of our humanity so social, emotional, physical and spiritual. That is something that really resonates with us as a family and our own values. (QA)
>
> So the tinana [body] side of it is really important but you’ve also got the taha wairua [spiritual] and spiritual side of it that incorporates mana and self dignity. I don’t know if that is in the F words, I’m not suggesting that it needs to be a separate pillar or category in the F-words, but I think through all of this, particularly with children of disability, your self esteem and your dignity is massive. So the concept of mana and maintaining mana is important. (QC)

One participant of Māori culture explained the relationship between wairua (spirituality), mana (self-esteem), and whanau (family).

> It’s a bit of a blend, it even falls under whanau [extended family] too. So it’s both. When your mana is acknowledged and respected, that helps with your wairua and the spiritual aspect. But mana generally is a major plank of mental well-being… Wairua is huge like that. Wai means water and rua means two, so the concept is and this comes back to your ancestors, but you are the outcome of your two parents and the two waters coming into you. So straight away you are looking beyond yourself and up to your parents and from their parents so you are part of a bigger collective. So mana I think mana fits in both wairua and hinengaro [psychological]. (QC)

Parents brought the concept back to very practical applications such as wheelchair access, toileting, and taking time to recharge as a caregiver.

> It would be for my child, fostering her self confidence and her identity as someone having a disability. That might include educating her peers and stuff like that. We have had to do a little bit of education with peers as kids would like to drive the power chair so we’ve had to do a bit of work in the background with the teachers to teach them that the power chair is just like my child’s legs and you wouldn’t take their legs so you don’t take the joystick for the power chair. (EQ)
>
> While the challenges are primarily physical, it leads on to her more general wellbeing, her mental wellbeing, her mana, her self esteem. Not being able to participate in activities and not being able to participate physically but also in her presence because she needs to go to the toilet or she’s got medical appointments or she’s having a… surgery, all of those things would have an impact on her mental well-being. (EQ)
>
> So in part of a week, then each of us needs to make sure that we are having some time to recharge, in whatever form that works for that person. And I guess that comes back to that kind of spiritual well being like, what is it? That’s yeah, what is it that’s underpinning each individual and the focus isn’t, or isn’t just on, on the child, the focus is on us as a collective. That, that well-being bit of us as a collective isn’t always necessarily interconnected. (QT)

One parent expressed how these concepts underpin wellbeing:

> I think the main thing is the wairua [spiritual] and the hinengaro, which is your mind and mental well-being, as these need to be somewhere in there, not necessarily separate topics but incorporated through a lot of the goals. (QC)

## Discussion

This study illustrates the burden experienced by families of children with developmental needs. This burden is exacerbated by expert centered care and alleviated by the FWLW approach. The holistic framework provided by the F-words coupled with professional coaching transformed the journey for families by giving them ownership of their goals. Our analysis shows the need for more recognition of connectedness, a term we use to encompass the concepts of spiritual connectedness and mental wellbeing.

Previous studies have shown the difficulties faced by families of children with disabilities (DePape & Lindsay, 2015; Graungaard et al., 2011; Oti-Boadi, 2017; Samsell et al., 2022). In a review of the literature on the experiences of parents of children with autism spectrum disorder, DePape and Lindsay reported that some parents feel overwhelmed, stressed, exhausted, and drained (DePape & Lindsay, 2015). Graungaard et al. noted that “initially parents spent much mental energy on coping with the shattered dreams and expectations as they became aware of the disabilities of their child” (Graungaard et al., 2011) and note that “the paralysing effect on coping abilities of negative feelings, such as fear, guilt and despair, were often reported by the parents” (Graungaard et al., 2011).

Boström et al. described the negative experiences of parents of children with intellectual disability, but also showed that positive experiences can balance the negative (Boström et al., 2010).

Our analysis showed the importance of empowering parents of children with developmental needs. DePape and Lindsay showed that some parents of children with autism spectrum disorder grew to become educators, therapists, and advocates at home, amongst their family and friends, and in the community (DePape & Lindsay, 2015). Empowered by knowledge and action, these parents could challenge information they received from others and develop a sense of control over their lives (DePape & Lindsay, 2015). Similarly, the parents in our study felt more empowered through the action of setting their own goals.

Centering power with parents and children aligns with the self-determination theory of human motivation. Self-determination theory is based on the premise that behavior originates from the self, and recognizes patient autonomy as one of three fundamental psychological needs required for motivation and goal achievement. (Deci & Ryan, 2012). The three psychological needs − autonomy, feelings of competence, and relatedness to others − should be fostered in order to motivate individuals to achieve their goals (Deci & Ryan, 2012). The FWLW increased autonomy by ensuring parent voice, and increased competence by taking a strengths-based approach. Relatedness was fostered within the FWLW by taking into account family and friends – “F-words” which ensure children and their family have robust social support systems. Thereby, the FWLW ensures that all three psychological needs are met for motivation and effective goal setting.

The psychological needs of autonomy, feelings of competence, and relatedness to others are not only important for achieving goals but for general coping as well. Some parents of children with developmental needs cope better than others (Graungaard et al., 2011) which can lead to an apparent paradox: people living full and meaningful lives in the face of significant disability (Albrecht & Devlieger, 1999; Larson, 1998). When people have more resources they are better equipped to cope (Fredrickson, 2004; Graungaard et al., 2011; Stifter et al., 2020). Resources include strong social relationships, protected family routines, taking action, and the normalization of circumstances, all of which fit with the FWLW approach in which families set goals that are important to them.

Coping through resource creation aligns with Fredrickson’s broaden and build theory of positive emotions. Fredrickson argued that positive emotions broaden peoples’ perspectives which helps build mental and physical resources (Fredrickson, 1998, 2004; Stifter et al., 2020). This in turn promotes wellbeing, although wellbeing is more than just positive emotions. Wellbeing can be thought of as a complex system in which individuals, institutions, and systems participate (Lomas et al., 2021). This fits with the rebalancing of power theme of the present study. Positive power comes from the interaction of the child, parents, and therapists. Interactions are both broad (holistic) and long (journeying). Thinking about wellbeing as a system also explains how rebalancing power is limited when some institutions, for example the education or medical systems, fail to participate in a holistic approach such as the FWLW because each part of the system plays its part in influencing wellbeing.

The F-words operationalize the ICF framework (Rosenbaum, 2022); in turn, the FWLW operationalize the F-words (Charles et al., 2020) through the use of coaching. Other frameworks have found coaching a way to put theory into practice. Social Determinant Theory researchers addressed the problem of translating their theoretical concepts into action (Patrick & Williams, 2012) through Motivational Interviewing (Miller & Rose, 2009) which, like Social Determinant Theory, has its roots in Roger’s client-centered empathy (Rogers, 1957, 1959). It is interesting that the key to operationalizing both approaches was through personal connection: coaching (Graham, 2020) in the FWLW and Motivational Interviewing for Social Determinant Theory (Vansteenkiste & Sheldon, 2006) in keeping with two of the themes from the present study: the rebalancing power theme resonates with autonomy and motivational shift (Patrick & Williams, 2012) while the connectedness theme resonates with empathy and connection characterized by coaching.

The connectedness theme pulled together the seemingly disparate concepts of mental health, sense of self, personal power, ancestry, and spirituality. We asked interviewees about their view on Māori concepts of health and heard about connections between the body, self, and the environment. In the Māori world-view, the sky, waters (oceans, rivers, wetlands), land, trees, animals, and people are all linked together (McNeill, 2017). Māori view health holistically: the physical, psychological, social, and spiritual all contribute to wellbeing. Most importantly, wellbeing is anchored in the environment (Durie, 1998; McNeill, 2017), particularly in relation to a sense of place (Panelli & Tipa, 2007). Similar holistic models of well-being exist amongst other Indigenous Peoples around the world (Redvers et al., 2022) including those in Peru (Izquierdo, 2005) and Canada (Beaudin, 2011).

Research shows that connectedness is an important determinant of health (K. C. Townsend & McWhirter, 2005). Connectedness encompasses a sense of self (intrapersonal connectedness) and social relations (interpersonal connectedness) (Sheilds, 1995). A sense of self relates to agency and empowerment (Curtis, 1992; Rude & Burnham, 1995), thereby the “connectedness” theme of the present paper encompasses the “re-balancing power” theme because connectedness is at least partially responsible for that power. A sense of self can also be experienced as interdependent and connected (Rude & Burnham, 1995). In understanding a sense of self, “an experiential sense of community and an awareness of a union with something larger than ourselves − that is, the nonlogical ‘connecting’ with the experience of others − would need to be valued as much as the awareness of our autonomy, agency, and uniqueness” (Curtis, 1992). This is where inner connectedness appears to be most important. Inner connectedness and sense of self may also relate to the sense of connection to ancestry and the environment, an important feature of many cultures. Furthermore, connectedness fits with Engel’s “continuum of natural systems” representation of the biopsychosocial model which describes a series of nested systems: individuals exist within family and social systems which in turn exist within the biosphere (Engel, 1981).

Caring for a child with developmental needs can benefit parents’ sense of connectedness by strengthening family bonds, bringing parents closer together in their relationships, altering worldviews, and bringing a sense of spiritual change (DePape & Lindsay, 2015). This can lead to an apparent paradox whereby people can live lives of joy, hope, and personal growth in the face of significant disability (Albrecht & Devlieger, 1999; Larson, 1998). Parents who can create meaning and experience positive emotions cope better than those who do not (Albrecht & Devlieger, 1999; Graungaard et al., 2011). Reminiscent of Carse’s “finite and infinite games” (Carse, 2019), connectedness may help frame the metrics of achievement, ability and disability as “finite games” while framing close relationships with other humans and nature as a timeless “infinite game” were participation, not winning, is what counts.

### Modeling

The themes from this study follow a temporal flow from overwhelming → power rebalance → connectedness. Families seemed to move from a sense of being overwhelmed to becoming rebalanced by taking agency of their care and moving forward to a sense of increasing connectedness. The themes from this study can be viewed as nested: the technical paradigm that leads to a sense of being overwhelmed can be conceptualized as smaller and (ideally) subordinate to a larger holistic paradigm whereby power is rebalanced back to the family. The holistic paradigm in turn sits within the connectedness theme as shown in Figure 1.

**Figure 1.**
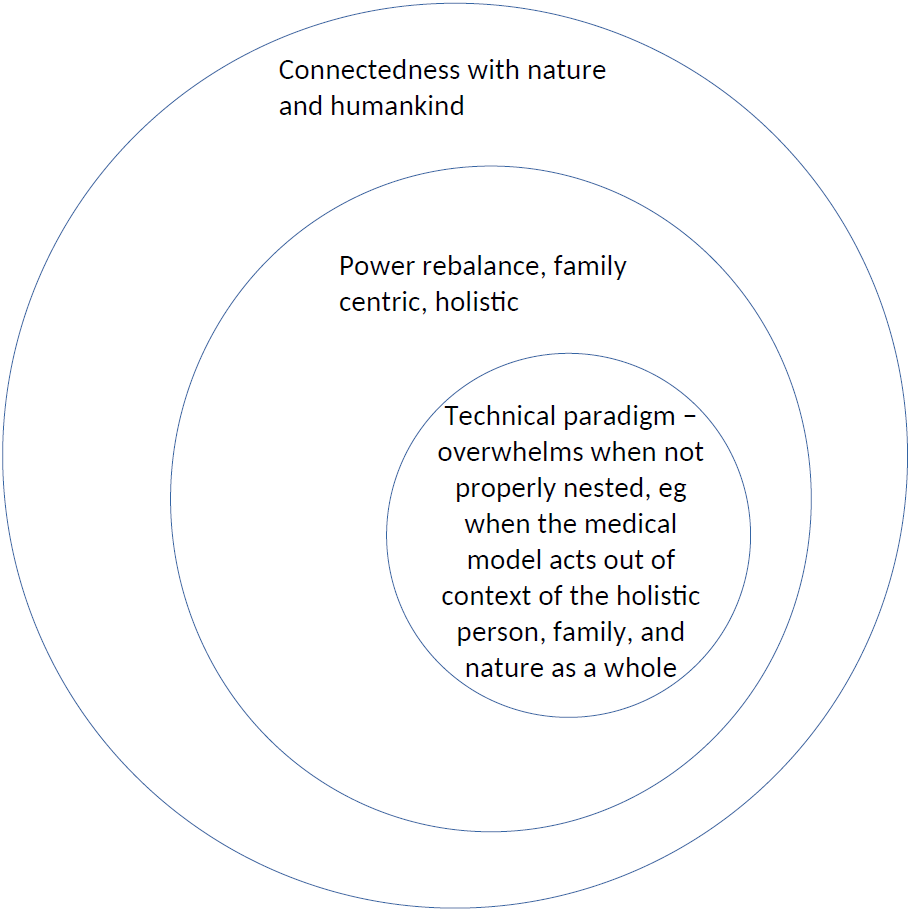
Nesting of themes.

Another dimension of connectedness which was not a theme in the present study is connection with nature. Numerous studies have shown a connection between well-being and connectedness with nature (Moll et al., 2022; Olivos & Clayton, 2017; Pritchard et al., 2020; Samus et al., 2022; Seers et al., 2022). This fits with the connection with the land in Durie’s hauora model (Durie, 1998), with spiritual connection with nature (the sky, ocean, land, trees, etc.) characteristics of traditional ecological knowledge from Māori indigenous worldview (Ihirangi, 2021) to Irish folk-law (Simon, 2000). One framework based on the Māori worldview is Rauora:

> The Rauora model depicts a worldview indigenous to Aotearoa; it centralises interconnection, collectivity, holistic wellbeing and intergenerational equity within a changing environmental dynamic. That changeable future is understood by first positioning the human existence within a sacredly interconnected world (Ihirangi, 2021).

Connectedness has practical implications for therapy. Townsend and McWhirter recommended that practitioners cultivate within themselves a strength-based attitude and a “nonjudgemental observational stance towards their clients’ connectedness” (K. C. Townsend & McWhirter, 2005). Cultivating one’s own sense of connectedness as a practitioner may be a better starting point than trying to append the concept to an existing model, for example, by adding another “F-word”. Connectedness could instead be conceptualized as moving through all the that we do in therapy.

We hope that readers whose child development experience differs from the present study will find some resonance (Tracy, 2010) with our findings. The “overwhelmed” theme may have transferability to parents of children with developmental needs in many other situations. The “power rebalance” theme may find naturalistic generalization to other settings where complex needs are being provided for, for example, chronic medical conditions, adult disability, etc. Finally, we hope that the “connectedness” theme may resonate across healthcare as an oft-ignored dimension of real importance in health and wellness.

### Limitations

This study used a critical realism approach. This was appropriate for a first look into the effect of the FWLW and also avoided the traps of ontic fallacy and epistemic fallacy that science tends to fall into, especially in the social sciences (Pilgrim, 2014). The number of interviewees was relatively small. Parents of children with developmental needs comprised most of the sample but two health professionals were also included, adding heterogeneity to the sample which according to Malterud *et al*. would indicate the need for a larger sample (Malterud et al., 2016). Further research is needed on health professionals’ views on holistic coaching models in other settings such as public health services or indigenous-based health services. Some themes were built on the discourses of a limited number of parents in the study. This study was also limited by not having performed member checking whereby the themes are discussed with the participants themselves for their feedback, something that could be performed in future studies.

## Conclusions

In conclusion, the journey through childhood disability is long and tough for families. Parents find the FWLW comprehensive, effective, and enjoyable. There is not only space for a holistic framework such as the FWLW in clinical practice, there is a need for it. The FWLW is empowering and motivating for children and families. If developed to be made more mainstream, it could positively impact many other children’s and families’ lives.

## Data Availability

All data produced in the present work are contained in the manuscript

## Acknowledgements

Thank you to Mercedes Mudgway for helping with transcription of two of the interviews and for your expert advice on themes.

## Appendix 1 Interview guide

### Guidance notes for the interviewers

Start the interview by establishing connection, telling a little about yourself and finding out something about your interviewee. Try to gather all the demographic information. Questions are a general guide—they do not all have to be covered. Ask questions appropriate to the focus of your interview. Use open-ended questions and “curious’ questions. Prompts are useful— “Can you tell me more about this?”, “What would that look like?” It is a conversation. Let them tell their story.

### Preliminaries

Mihi: Share information about yourself, consider sharing your pepeha, listen and learn about your interviewee, where they are from and if appropriate their pepeha.

Karakia: If appropriate, invite a karakia or ask permission to say a karakia.

### Korero/discussion

#### Introduction

Thank you for agreeing to participate in this interview. I am interviewing you to find out your experiences and ideas about hauora and the Fav-words Life Wheel. There are no right or wrong answers to any questions, we are interested in your story, experiences, thoughts and ideas. Participation in this interview is voluntary. The interview will probably take about 30 – 60 minutes. With your permission I would like to record this interview so as to not miss any comments. The recording will be transcribed and kept confidential. After I have transcribed it I will send you a copy so that you can check it and add/correct anything you like. Any information from your interview that appears in our report will not identify you. You may decline the interview or stop at any time.Do you have any questions?

#### Interview

- Information and consent
  - Explanation—information sheet (if performed on teleconferencing, email information sheet and consent form before the interview)
  - Consent form signed (if performed on teleconferencing, ask participant to email the signed consent form back)
  - Begin recording only after fully informed consent has been completed and returned by participants to the researchers
- Demographic information
  ∘ Age bracket
    ▪ <25
    ▪ 25 – 34
    ▪ 35 – 44
    ▪ 45 – 54
    ▪ 55 – 64
    ▪ 65+
  ∘ Gender
    ▪ Female
    ▪ Male
    ▪ Gender diverse
    ▪ Prefer not to say
  ∘ Ethic group(s) – allow multiple
    ▪ Māori
    ▪ European
    ▪ Pacific peoples
    ▪ Asian
    ▪ Middle Eastern/Latin American/African
    ▪ New Zealander
    ▪ Other ethnicity
  ∘ Professional training
  ∘ Role at present
- Interview questions section 1 – Fav-words
  ∘ What is your experience with the Fav-Words Life Wheel?
  ∘ How long have you been using it?
  ∘ What difference has it made? Examples?
  ∘ Ask them to rate their experience of using the Fav-words life wheel out of 10
  ∘ What settings do you find you use the Fav-Words Life Wheel in most? Ie just in a clinical setting or do you often use it at home as well?
  ∘ Have you used any other disability frameworks apart from the Fav-words life wheel? Could you share a little bit about them? Did you find there was an adjustment period to using the Fav-words model or was it quite easy to use?
  ∘ How different do you think it would have been if your therapist had used a different approach?
  ∘ How would you recommend scoring for each aspect of the wheel? IE does 1-10 work best or is there another way you would recommend?
  ∘ What has your experience been with the coaching side?
    ▪ The Fav words life wheel chooses to follow a coaching framework with the aim of allowing clinicians to work in partnership with parents and families, rather than simply telling them what to do.
  ∘ Do you have any improvements you’d like to see with the Fav-words model?
- Māori health model
  ∘ Can you tell me about your understanding or experience of the Māori cultural approach to health and wellness?
    Hauora Māori or Te Whare Tapa Whā This model takes into account the four pillars of health which are taha tinana (physical health), Taha wairua (spiritual health), Taha whānau (family health) and Taha hinengaro (mental health). The idea is that these are four pillars of a wharenui and when one pillar is weakened then a person becomes subsequently unwell.
  ∘ So taking into account the aspects of Māori health that we just discussed, how do you think the Fav-Words Life Wheel would look/fit in a Māori health approach to health and wellness?
  ∘ What do you think are the most important aspects of health and wellness from a Māori perspective? What are the key words for these?
  ∘ Please tell me about your experience with talking together/korero in health and wellness from a Māori perspective? For example, have you been a part of a clinical setting where there has been a pepha given or a karakia given?
    ▪ Do you think that you would like more Māori cultural values in health and wellness to be brought into clinical experiences that you have had?
    ▪ Would you like to see a pepeha or a karakia given in some situations?
  ∘ If you had to change the Fav-Words life wheel model in some way to better fit with Māori health and wellness values, how would you do this?
- Māori culture in staff briefings in operating theatres1
  ∘ How do you think we could better integrate Māori culture into clinical practice in the operating theatre for staff

#### Wrap up

Anything else they would like to add? So we will write up the transcript of the interview and send it to you within the next week. Please feel free to read over it and if you want to change or edit any of your answers then just let us know. Thanks.

## Appendix 2 Quality checklist

Braun and Clarke’s 15-point checklist of criteria for good thematic analysis (Braun & Clarke, 2006) and our quality self-assessment.

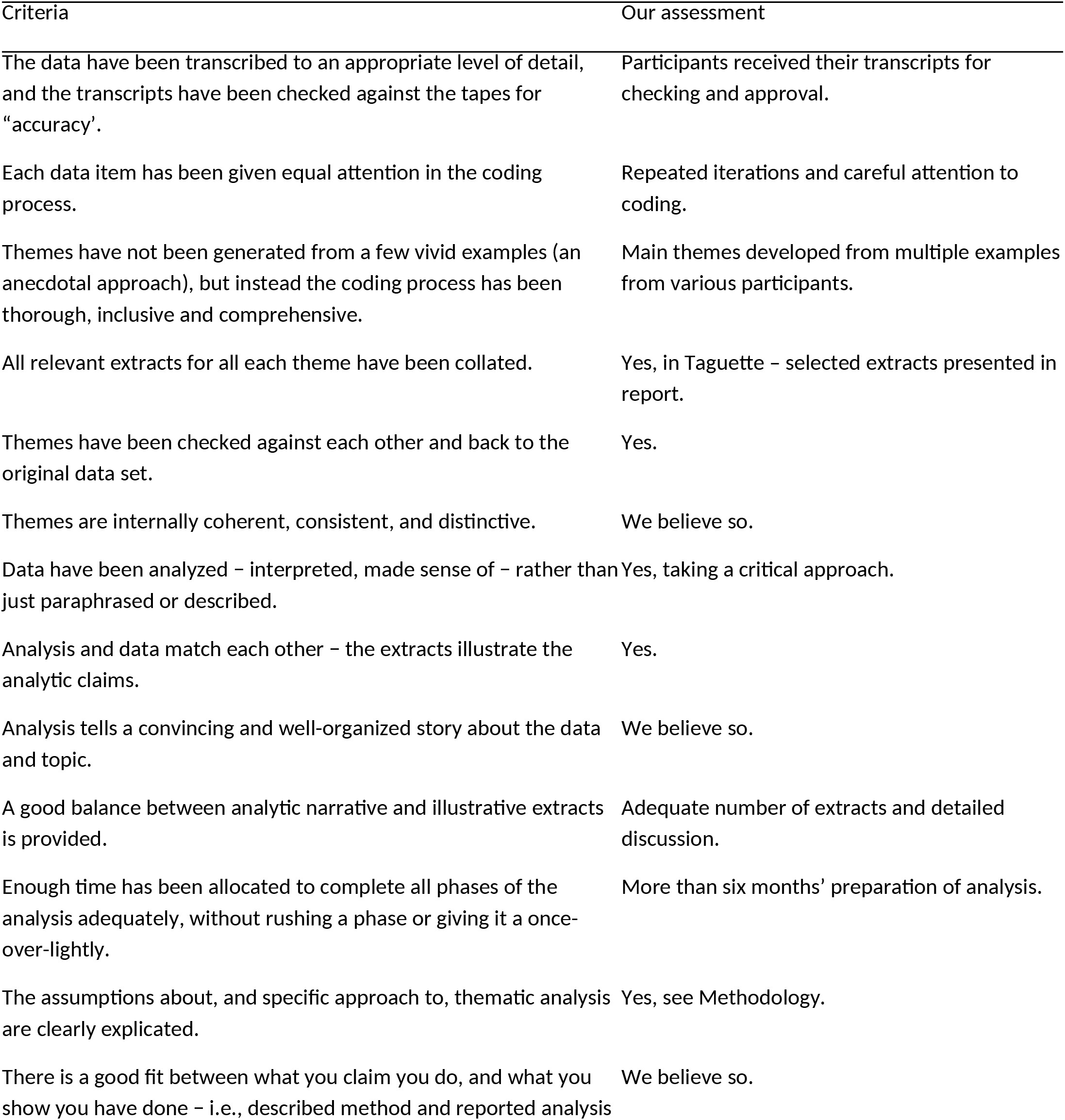
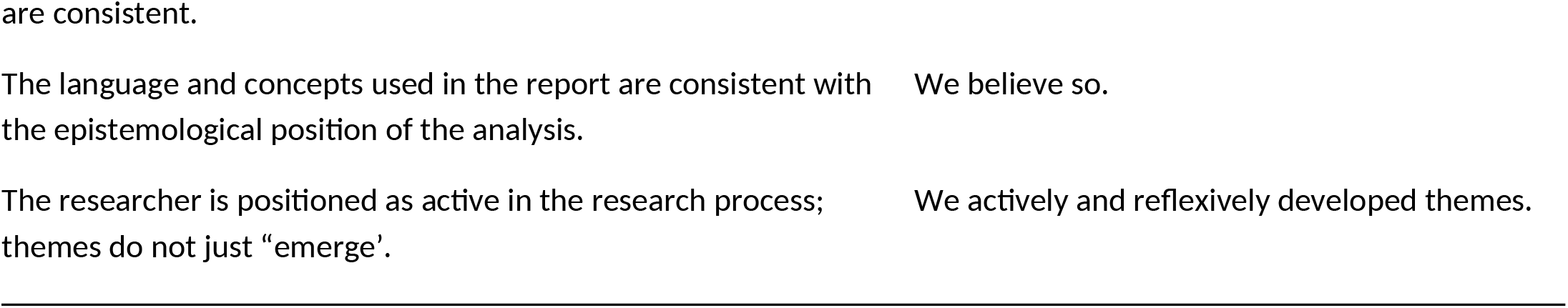

## Appendix 3 Conceptual depth scale

Nelson’s 5-point conceptual depth scale (Nelson, 2017) and our quality self-assessment.

| Criteria | Our assessment |
| --- | --- |
| Range (e.g. frequency and variety of codes, multiplicity of data sources) | We had a fairly extensive variety of code which we worked and re-worked and went through data multiple times |
| Complexity (e.g. coding trees, positional maps, matrices) | Conceptual diagrams are presented in Figure 1 |
| Subtlety (e.g. memos; social worlds diagrams) | We looked for subtle meanings behind some common terms particularly as they related to our theme of connectedness |
| Resonance (literature) | We have drawn on literature and models in the fields of disability, Māori health, and the environment, especially in the third theme |
| Validity (e.g. applicability test) | The findings should resonate with families of children with developmental needs as well as with care providers who might see this example of family centered care is applicable in other healthcare settings |

